# Plasma pTau181 reveals a pathological signature that predicts cognitive outcomes in Lewy body disease

**DOI:** 10.1101/2024.04.12.24305710

**Authors:** Carla Abdelnour, Christina B Young, Marian Shahid-Besanti, Alena Smith, Edward N. Wilson, Javier Ramos Benitez, Hillary Vossler, Melanie J. Plastini, Joseph R. Winer, Geoffrey A. Kerchner, Brenna Cholerton, Katrin I. Andreasson, Victor W. Henderson, Maya Yutsis, Thomas J Montine, Lu Tian, Elizabeth C. Mormino, Kathleen L. Poston

## Abstract

Lewy body disease (LBD) often co-exists with Alzheimer’s disease (AD), influencing disease progression, cognitive decline, and neurodegeneration. This study aims to determine whether plasma phosphorylated-Tau181 (pTau181) could be used as diagnostic biomarker of concurrent Alzheimer’s disease neuropathologic change (ADNC) or amyloidosis alone, as well as a prognostic, monitoring, and susceptibility/risk biomarker for clinical outcomes in LBD. Our sample comprised 565 Stanford research participants: 94 LBD with normal cognition, 83 LBD with abnormal cognition, 114 AD, and 274 who were cognitively normal. We measured plasma pTau181 levels with the Lumipulse *G* platform. Diagnostic accuracy for concurrent ADNC and amyloidosis was assessed with receiver-operating-characteristic curves in a subset of participants with CSF pTau181/Aβ42, and CSF Aβ42/Aβ40 or amyloid-β PET, respectively. We used linear mixed effects models to examine the associations between baseline and longitudinal plasma pTau181 levels and clinical outcomes. Plasma pTau181 predicted concurrent ADNC and amyloidosis in LBD with abnormal cognition with 87% and 72% accuracy, respectively. In the LBD with abnormal cognition, higher baseline plasma pTau181 was associated with worse baseline MoCA and CDR-SB, as well as accelerated decline in CDR-SB. Additionally, in this group rapid increases in plasma pTau181 over 3 years predicted a faster decline in CDR-SB and memory. In LBD with normal cognition, there was no association between baseline or longitudinal plasma pTau181 levels and clinical outcomes; however, elevated pTau181 at baseline increased the risk of conversion to cognitive impairment. These findings suggest that plasma pTau181 is a promising biomarker for concurrent ADNC and amyloidosis in LBD. Furthermore, plasma pTau181 holds potential as a prognostic, monitoring, and susceptibility/risk biomarker, predicting disease progression in LBD.

## INTRODUCTION

Neurodegenerative diseases are characterized by the abnormal deposition of multiple proteins, referred to as proteinopathies. Understanding whether interplay among proteinopathies impacts clinical outcomes is crucial to understand the heterogeneity in clinical presentation and rates of progression, and to guide drug development^1^.

Alzheimeŕs disease (AD) and Lewy body disease (LBD) are the two most prevalent neurodegenerative diseases. LBD includes Parkinsońs disease (PD) and dementia with Lewy bodies (DLB), and is characterized by the abnormal deposition of α-synuclein. In contrast, AD is associated with the deposition of amyloid-β and tau. These proteinopathies often coexist, with over 50% of clinically diagnosed LBD patients exhibiting concurrent Alzheimer’s disease neuropathologic change (ADNC) at autopsy^2^. The presence of concurrent ADNC in clinical LBD has an impact on disease progression, clinical phenotype, and brain atrophy patterns^3–8^.

Biomarkers enable in vivo investigation of brain proteinopathies. Emerging blood-based biomarkers like plasma phosphorylated-Tau181 (pTau181) have shown promise in detecting ADNC (i.e., diagnostic biomarker), predicting disease progression (i.e., prognostic biomarker), tracking changes over time (i.e.; monitoring biomarker), and assessing the risk of developing AD dementia (i.e., susceptibility/risk biomarker)^9–13^.

However, research on plasma pTau181 in LBD remains limited. Few studies have investigated its potential as a diagnostic biomarker for ADNC (i.e. amyloid-β plus tau) or amyloidosis alone, or its roles as a prognostic, monitoring, or susceptibility/risk biomarker. While it shows promise in detecting concurrent ADNC and amyloidosis in LBD patients with cognitive impairment^14–16^; its diagnostic utility in LBD patients with normal cognition is underexplored. Regarding its prognostic and monitoring value, one study associated higher baseline plasma pTau181 levels with worse cognition over time in DLB^14^, whereas another found no relationship between longitudinal changes in its levels with cognitive decline in mild cognitive impairment (MCI) due to DLB^16^. Moreover, no studies have investigated its prognostic value on daily functioning or neuropsychological performance in LBD. Lastly, as a susceptibility/risk biomarker, one study found no predictive value for dementia conversion in PD patients with normal cognition or MCI^17^. Importantly, no studies have explored its potential use for identifying LBD patients with normal cognition likely to develop MCI or dementia.

Further research in LBD is warranted to examine the relationship between longitudinal changes in ADNC biomarkers and clinical outcomes like global cognition, daily functioning, and neuropsychological performance. The prospective use of plasma pTau181 as a monitoring biomarker is relevant for understanding the dynamic progression of neurodegenerative diseases, which will inform patient management and prognosis, and clinical trial designs, serving as roadmap to precision medicine.

In this study, our objectives were to determine whether plasma pTau181 in people with LBD could serve as a diagnostic biomarker for ADNC and amyloidosis, predict baseline and longitudinal clinical outcomes (i.e.; prognostic and monitoring biomarker), and determine its potential as a predictor of cognitive impairment conversion in LBD patients with normal cognition (i.e.; susceptibility/risk biomarker). We hypothesized that plasma pTau181 levels in people with LBD will serve as a diagnostic biomarker for ADNC and amyloidosis, associate with clinical outcomes, and predict the conversion to cognitive impairment in those with normal cognition.

## METHODS

### 1.1. Research participants

We selected research participants with available plasma from the Iqbal Farrukh and Asad Jamal Stanford Alzheimer’s Disease Research Center (ADRC) and Pacific Udall Center (PUC)^18^ cohorts. Diagnostic procedures are described in detail elsewhere^19^.

Briefly, each participant underwent a comprehensive evaluation, encompassing clinical history, physical and neurological examination, and neuropsychological assessments. Diagnoses were established during multidisciplinary consensus meetings involving at least two neurologists, a clinical neuropsychologist, and other study personnel. LBD with normal cognition (LBD-nlCog) was established in participants diagnosed with PD according the UK Brain Biobank criteria^20^ who had no objective impairment on comprehensive neuropsychological testing. LBD with abnormal cognition (LBD-abnlCog) was established if the participant had diagnosis of 1) MCI due to PD^21^ or prodromal DLB^22^ (probable or possible), or 2) dementia due to PD^23^ or probable DLB^24^, according to published criteria. The AD group included participants diagnosed with MCI or dementia due to AD according to NIH Alzheimer’s Disease Diagnostic Guidelines^25^.

Cognitively normal (CN) participants were older adults with no parkinsonian symptoms, normal neurological examination, and performance on comprehensive neuropsychological testing that was normal for age and sex. We excluded participants with neurodegenerative diseases other than LBD or AD, large vessel stroke by history or magnetic resonance imaging (MRI), major psychiatric disorder, untreated severe mood related-disorder, toxic-metabolic encephalopathy, renal or hepatic disorders, autosomal dominant gene mutations for AD, learning disability, and cognitive impairment with unknown underlying etiology.

From a total of 633 participants, 565 individuals met the above inclusion criteria: 94 LBD-nlCog, 83 LBD-abnlCog, 114 AD, and 274 CN **(Supplementary Figure 1)**. A subset of 270 participants underwent yearly blood sampling. Within this subset, baseline data were available for all 270 individuals, with 254 having data at year 1, 55 at year 2, and 69 at year 3.

### 1. 2. Clinical and neuropsychological outcomes

Daily functioning was measured with the Clinical Dementia Rating sum of boxes (CDR®-SB)^26^, and motor function with the Movement Disorders Society-Unified Parkinson’s Disease Rating Scale (MDS-UPDRS) Part III^27^. Global cognition was measured with the Montreal Cognitive Assessment (MoCA)^28^ in 475 participants, and with the Mini-mental State Examination (MMSE)^29^ in 90 participants: 64 CN, 24 AD, and 2 LBD. MMSE scores were converted to MoCA scores according to age, sex, and years of education based on Monsell et al^30^.

For analyzing neuropsychological performance, we selected ADRC participants who completed the National Alzheimer’s Coordinating Center (NACC) Uniform Data Set version 3 (UDS-3) neuropsychological battery described elsewere^31^, in addition to supplemental cognitive tests for memory, executive and visuospatial functions. For each neuropsychological test, z-scores were calculated using means and standard deviations of baseline visit from all CN participants (i.e., not only participants with biomarker data) in the ADRC. The z-scores were then used to create domain-specific composite scores by averaging all relevant z-scores for the domain. The memory composite consisted of Craft Story 21 delayed recall, Hopkins Verbal Learning Test–Revised delayed recall^32^, Benson Complex Figure delayed recall, and Free and Cued Selective Reminding Test delayed free recall^33^ (required at least 3 out of 4 completed tests); the executive functioning composite consisted of Trails B, Victoria Stroop color/word time^34^, WAIS III Digit Coding Subtest, Clock Drawing Test, phonemic fluency, and semantic fluency (required at least 4 out of 6 completed tests); the attention/working memory/processing speed composite consisted of Trails A, Number Span Forward, Number Span Backward, Victoria Stroop color and word reading times separately^34^, and Letter-Number Sequencing^35^ (required at least 4 out of 6 completed tests); visuospatial function composite consisted of Judgment of Line Orientation^36^, Clock Drawing Test copy, and Benson Complex Figure copy (required at least 2 out of 3 completed tests); and the language score consisted of Multilingual Naming Test (MINT).

The levodopa equivalent daily dose (LEDD) was calculated according to published criteria^37^, and LBD participants taking dopamine replacement therapy completed cognitive testing in the on medication state.

All baseline and longitudinal test scores were obtained within a 6-month window of plasma collection. **Supplementary tables 1 and 2** show the number of participants with longitudinal clinical outcomes.

### 1. 3. Biomarkers collection and analysis: plasma, CSF and amyloid PET

Plasma pTau181 levels were determined using a modified version of Lumipulse *G* CSF pTau181 assay (Cat. # 231654, Fujirebio Diagnostics, US, Malvern, PA) with the LUMIPULSE *G*1200 instrument, as previously detailed^10^. The Lumipulse *G* plasma pTau181 assay employs a combination of antibodies based on INNOTEST assay, that targets tau epitopes near Thr181. This combination includes the capture antibody AT270, and detection antibodies HT7 and BT2^38^. To prepare plasma samples, we thawed them on wet ice, followed by a 5-minute centrifugation at 4°C and 500 × g.

Subsequently, the samples were loaded onto the fully automated LUMIPULSE *G*1200 instrument. To reduce the potential for non-specific binding, we pre-treated plasma samples with a heterophilic blocking reagent (200 μg/ml, Scantibodies Inc., Santee, CA)^39^. We assessed individual-level variability using 6 independent plasma aliquots and a different batch of reagents one year later, demonstrating high test-retest reliability (Pearson’s r = 0.98). All plasma samples from our current study fell within the quantifiable range, which spanned from 0.16 to 10.43 pg/ml.

CSF was obtained through the following steps: 1) a lumbar puncture was performed at the L4-L5 or L5-S1 interspace using a 20-22 G spinal needle; 2) CSF was collected in externally threaded Thermo Scientific™ Nalgene™ General Long-Term Storage Cryogenic Tubes; and 3) CSF samples were stored in aliquots at −80°C until analysis, with a maximum of two freeze-thaw cycles.

CSF biomarkers, including Aβ42, Aβ40, pTau181, and total tau, were quantified using the LUMIPULSE *G*1200 instrument by the Stanford ADRC Biomarker Core. The cut-points for CSF biomarkers, determined using the Youden method to optimize sensitivity and specificity to discriminate clinically-defined AD from CN^10^, were as follows: abnormal Aβ42/Aβ40 ratio < 0.09, abnormal pTau181 > 87.34 pg/mL, abnormal total tau > 592.26 pg/mL, and abnormal pTau181/Aβ42 ratio > 0.13.

Amyloid-β PET imaging was performed with ^18^F-florbetaben in a PET/MRI scanner (Signa 3 T, GE Healthcare) at the Richard M. Lucas Center for Imaging at Stanford University. Data on emissions were gathered from 90-110 minutes after injection of 8.1 miliCuries of ^18^F-florbetaben. PET data were reconstructed into 5-minute intervals using conventional techniques. Zero-Time-of-Eco (ZTE) and Dixon-based MR imaging was used for MR attenuation correction (MRAC), and the data from 5-minute frames were realigned and combined.

Biomarker evidence of ADNC was established with abnormal CSF pTau181/Aβ42 ratio. Biomarker evidence of amyloidosis was established with abnormal CSF Aβ42/Aβ40 ratio or visual assessments of amyloid-β PET scans. For the latter, a scan was considered positive with a consensus rating by at least two out of three experienced radiologists, blinded to the clinical diagnosis. In cases of discordance between CSF Aβ42/Aβ40 ratio and amyloid-β PET imaging, preference was given to the CSF ratio result.

All biomarker processing (plasma, CSF and amyloid-β PET imaging) was performed blinded to clinical information. CSF samples used in the analyses were obtained within 6 months of plasma collection (mean 2.6 weeks, SD: 4.7), and amyloid-β PET imaging was obtained within 1 year of plasma collection (mean 10.95 weeks, SD: 9.9).

From the 565 research participants who met inclusion criteria, 245 had AD CSF biomarkers results and/or amyloid-β PET imaging available: 115 CN, 47 LBD-nlCog, 47 LBD-abnlCog, and 36 AD. Two-hundred and one participants had AD CSF biomarkers, 73 had amyloid-β PET imaging, and 29 had both.

### 1. 4. Statistical analysis

Due to the skewed distribution of plasma pTau181 levels, we performed a log10-transformation on their raw values. We used mean (standard deviation) to summarize approximately normally distributed continuous measures, median (range) to summarize non-normally distributed continuous measures, and counts (percentage) to summarize categorical variables.

To determine differences between diagnostic groups, we used either one-way ANOVA or the Kruskal-Wallis test, as appropriate. Subsequently, we performed post hoc pair-wise comparisons using t-test, and applied the Bonferroni correction to account for multiple testing. For categorical variables, we used either the chi-square test or Fisher’s exact test, depending on observed proportions.

We used the Receiver Operating Characteristic (ROC) curve and the area under the curve (AUC) to measure the accuracy of plasma pTau181 in distinguishing LBD-nlCog and LBD-abnlCog participants with and without concomitant ADNC or amyloidosis. In addition, we selected the optimum cut point used in a diagnostic test by maximizing the Youden J Index in the ROC curve.

We used multiple linear regression models to determine the association of the baseline plasma pTau181, diagnostic group, and their interaction with baseline clinical outcomes including global cognition (MoCA), daily functioning (CDR-SB), and neuropsychological performance, after adjusting for age and sex.

We then used linear mixed effects models for repeated measures (MMRM) to determine the associations of baseline and longitudinal changes in plasma pTau181 with the three-year decline in clinical outcomes of interest. In the first set of MMRM regression analyses, we included baseline plasma pTau181, diagnostic group, age at blood draw, sex, and their interaction with time as independent variables. In the second set of MMRM regression analyses, we first estimate the average slope in plasma pTau181 during 3-year of follow-up for each patient and additionally included this slope as an independent variable of interest in the MMRM regression analysis. The regression models included subject-specific random intercepts and slopes to account for the within-subject correlations. The MMRM regression yields valid inference results under the missing at random assumptions allowing missing longitudinal cognitive functional measures in some study participants. For regression models investigating global cognition and neuropsychological performance, we also included years of education as an independent variable.

Finally, we used Cox proportional hazard multiple regression model to predict the risk of conversion to cognitive impairment in the LBD-nlCog and CN groups using the baseline plasma pTau181 levels, age, and sex as predictors. Participants from both groups were categorized into those with normal and abnormal plasma pTau181 levels. This categorization was determined using the Youden J Index method in ROC curve analysis for distinguishing LBD-nlCog Aβ+ from Aβ-individuals. We then estimated the adjusted hazard ratios (HR) and 95% confidence intervals. Conversion to cognitive impairment was defined as progression from normal cognition to diagnosis MCI or dementia during follow-up.

All statistical analyses were performed with R statistical software (R Foundation for Statistical Computing, http://www-R-project.org) version 4.3.0, and IBM SPSS version 26. A two-sided p-value ≤0.05 was deemed statistically significant.

### 1. 5. Ethical considerations

The Institutional Review Board of Stanford University granted approval for the study protocols. All participants or their legally authorized representatives provided written informed consent for participation, according to the Declaration of Helsinki.

## RESULTS

### 1.1 Participants characteristics

Detailed description of the baseline characteristics of the participants is presented in **Table 1**. The LBD-nlCog group was younger than the CN (p<0.001) and AD groups (p<0.001). No significant differences in age were observed between the LBD-abnlCog group and the CN and AD groups. Both LBD subgroups (normal and abnormal cognition) were mostly male (51.1% and 72.3%, respectively). There was no difference in years of education, race, and ethnicity between diagnostic groups.

**Table 1:**
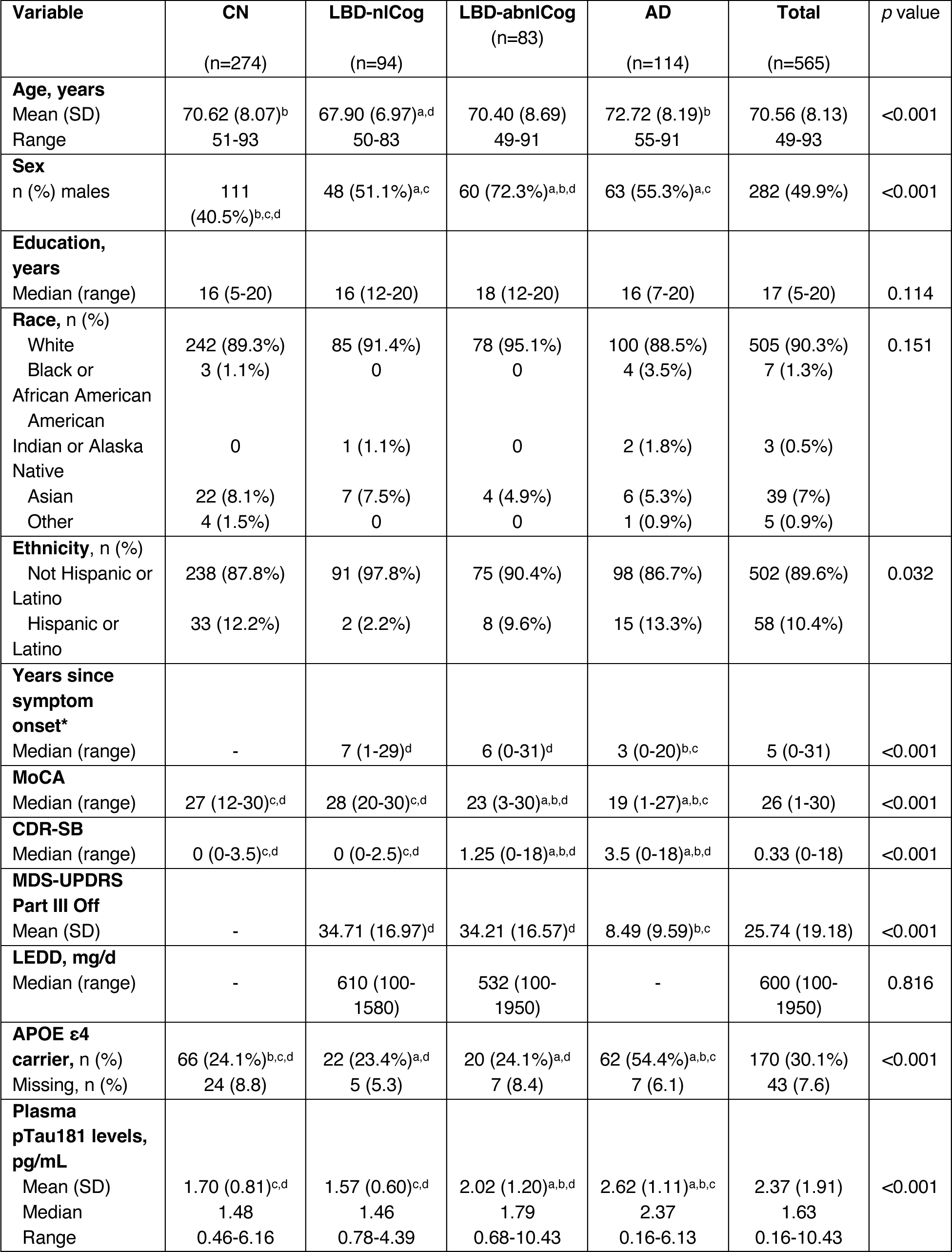
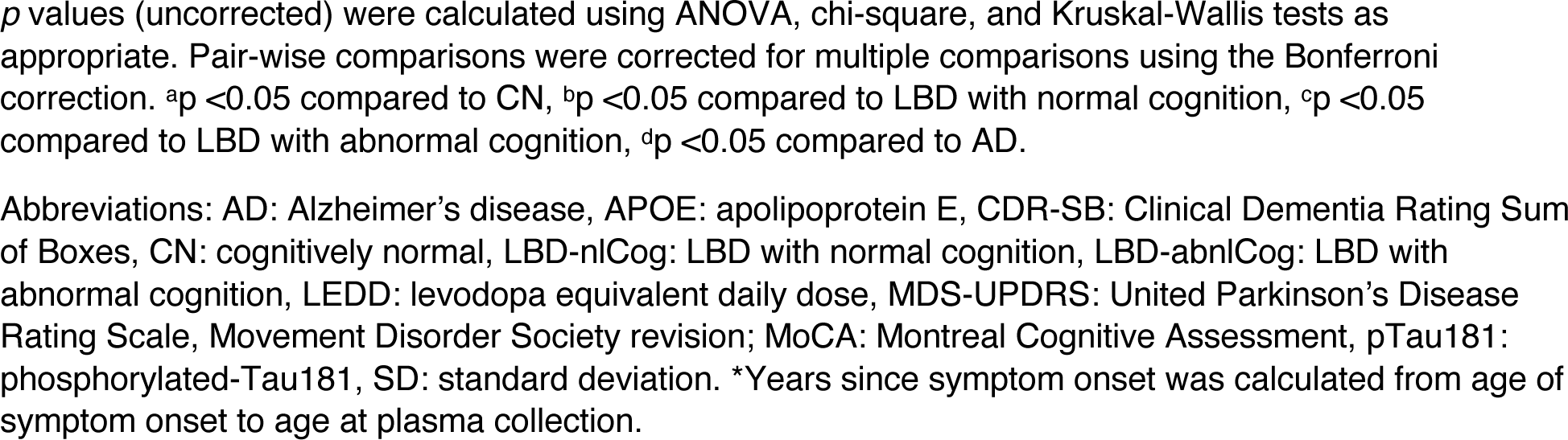
Characteristics of research participants.

Regarding clinical characteristics, the LBD-abnlCog group had significantly worse MoCA and CDR-SB scores compared to both the CN (p<0.001) and LBD-nlCog groups (p<0.001). However, their MoCA and CDR-SB scores were better than those of the AD group (p<0.001). There was no difference in MDS-UPDRS Part III scores, LEDD, or percentage of APOE ε4 carriers among LBD subgroups.

### 1. 2. Plasma pTau181 levels across groups

Participants in the LBD-abnlCog group showed higher plasma pTau181 at baseline compared to both LBD-nlCog (p<0.001) and CN groups (p<0.001), but their levels were lower than those observed in the AD group (p<0.001) **(Figure 1A)**. Notably, there was no difference in plasma pTau181 levels between LBD-nlCog and CN participants.

**Figure 1:**
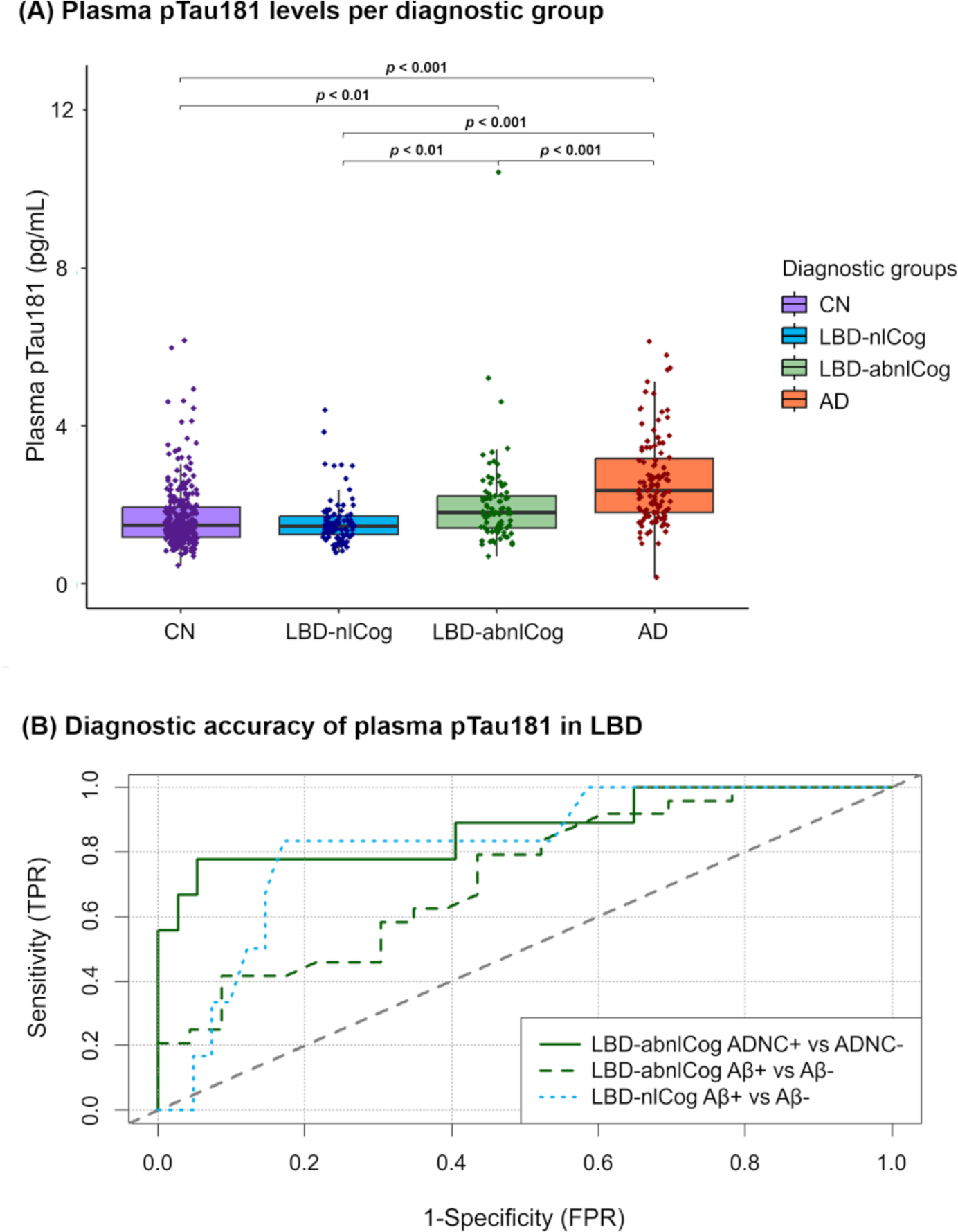
Plasma pTau181 as a diagnostic biomarker in Lewy body disease. **A)** Comparison of plasma pTau181 levels among diagnostic groups: CN (purple), LBD-nlCog (blue), LBD-abnlCog (green), and AD (orange). Differences were analyzed using log-transformed plasma values with ANOVA and corrected for multiple comparisons with the Bonferroni method. **B)** ROC curve analysis of plasma pTau181 levels in distinguishing: LBD-abnlCog ADNC+ (19.1%) vs ADNC-(80.9%) (continuous green line), LBD-abnlCog Aβ+ (51.1%) vs Aβ-(48.9%) (dashed green line), and LBD-nlCog Aβ+ (12.8%) vs Aβ-(87.2%) (dotted blue line). Abbreviations: Aβ: amyloid-β, AD: Alzheimer’s disease, ANOVA: analysis of variance, CN: cognitively normal, FPR: false positive rate, LBD-abnlCog: Lewy body disease with abnormal cognition, LBD-nlCog: Lewy body disease with normal cognition, pTau181: phosphorylated-Tau181, TPR: true positive rate.

In a sensitivity analysis, we excluded one outlier identified through visual inspection from the LBD-abnlCog group with a baseline plasma pTau181 level of 10.43 pg/mL. After removing this subject, the results remained unchanged.

### 1. 3. Plasma pTau181 as a diagnostic biomarker for ADNC and amyloidosis in LBD

Detailed description of the 245 participants with available AD CSF or amyloid-β PET biomarkers are available in **Supplementary Table 3**. This subgroup demonstrated no age or sex difference across the diagnostic categories, in contrast to the complete cohort (described in **Table 1**). However, with regards other demographic or clinical characteristics, this subgroup mirrored the complete cohort.

We analyzed whether plasma pTau181 levels could distinguish between LBD participants with ADNC (ADNC+) from those without ADNC (ADNC-), measured with the CSF pTau181/Aβ42 ratio. In the LBD-abnlCog group, we found that plasma pTau181 distinguished LBD-abnlCog CSF ADNC+ (19.1%) from ADNC-(80.9%) with an AUC= 0.87 (95% CI: 0.72-1.00), Positive Predictive Value (PPV)= 77.78%, and Negative Predictive Value (NPV)= 94.59% **(Figure 1B)**. None of the LBD-nlCog had an abnormal CSF pTau181/Aβ42 ratio.

Similarly, we analyzed whether plasma pTau181 could distinguish between LBD participants with amyloidosis (Aβ+) from those without amyloidosis (Aβ-), as measured either by abnormal CSF Aβ42/Aβ40 ratio or positive amyloid PET. In the LBD-abnlCog group, we found that plasma pTau181 distinguished Aβ+ (51.1%) from Aβ-(48.9%) with an AUC= 0.72 (95% CI 0.58-0.87), PPV= 65.52%, and NPV= 72.22% **(Figure 1B)**.

Moreover, in the LBD-nlCog group, plasma pTau181 distinguished Aβ+ (12.8%) from Aβ-(87.2%) with an AUC=0.82 (95% CI: 0.64-0.99), PPV= 41.67%, and NPV= 97.14% (Figure 1B).

In a sensitivity analysis, excluding the previously mentioned outlier from the LBD-abnlCog group did not change the results from the ROC model distinguishing CSF ADNC+ from ADNC-and distinguishing Aβ+ from Aβ-participants.

### 1. 4. Plasma pTau181 as a prognostic biomarker in LBD

In the LBD-abnlCog group, baseline plasma pTau181 levels were associated with worse baseline MoCA (β= −8.86 (SE= 2.34), p<0.01) and CDR-SB scores (β= 9.63 (SE= 1.61), p<0.001). However, there was no association between baseline plasma pTau181 levels with baseline neuropsychological performance. This might result from the fact that MoCA and CDR-SB are global measures, capturing overall cognitive and functional impairment, whereas the performance across different cognitive domains in people with LBD can present greater variability at baseline. Over a 3-year follow-up period, baseline plasma pTau181 levels were associated with a more rapid decline in CDR-SB (β= 3.20 (SE= 0.63), p<0.001), but not with longitudinal changes in MoCA or neuropsychological performance. These findings remained unchanged even upon exclusion of the previously mentioned outlier within this diagnostic group.

In the LBD-nlCog and CN groups, there were no associations between baseline plasma pTau181 levels and MoCA, CDR-SB or neuropsychological performance either at baseline or after 4 years of follow-up.

In the AD group, baseline plasma pTau181 levels were associated with worse baseline scores in MoCA (β= −12.32 (SE= 2.00), p<0.001), CDR-SB (β= 8.22 (SE= 0.91), p<0.001), memory (β= −1.45 (SE= 0.46), p<0.01), executive function (β= −1.74 (SE= 0.60), p<0.01), and language scores (β= −2.67 (SE= 0.84), p<0.01), but not with attention/working memory/processing speed or visuospatial function. After a 3-year follow-up period, baseline plasma pTau181 levels were associated with faster decline in MoCA (β= −3.22 (SE= 1.00), p<0.01), CDR-SB (β= 3.10 (SE= 0.44), p<0.001), and executive function (β= −0.98 (SE= 0.33), p<0.01), but not with longitudinal changes in memory, attention/working memory/processing speed, visuospatial function, or language.

### 1. 5. Plasma pTau181 as a monitoring biomarker in LBD

In the LBD-abnlCog group, a faster increase in plasma pTau181 levels over 3 years was associated with accelerated declines in CDR-SB (β= 1.97 (SE=0.80), p<0.05), and memory composite scores (β= 1.31 (SE=0.43), p<0.01). A trend association emerged between this faster increase in plasma pTau181 and executive composite score (β= - 0.70 (SE=0.38), p=0.06) as well as attention/working memory/processing speed composite score (β= −0.54 (SE=0.30), p=0.07). There was no association with longitudinal changes in MoCA, visuospatial function or language.

In contrast, in the LBD-nlCog group, there was no association between changes in plasma pTau181 levels up from baseline to 3 years of follow-up with longitudinal changes in MoCA, CDR-SB, or neuropsychological performance.

In the AD group, a faster increase in plasma pTau181 from baseline to 3 years of follow-up was associated with faster decline in MoCA (β= −5.55 (SE=1.26), p<0.001), CDR-SB (β= 2.65 (SE=0.40), p<0.001), and language scores (β= −1.54 (SE=0.67), p<0.05).

Finally, in the CN group, faster increases in plasma pTau181 levels from baseline up to 3 years of follow-up were associated with faster decline in attention/working memory/processing speed composite scores (β= −0.16 (SE=0.07), p<0.05).

**Table 2** and **supplementary figure 2** summarizes the associations between the 3-year change of plasma pTau181 levels and longitudinal clinical outcomes.

**Table 2:** Associations between 3-years change in plasma pTau181 levels and longitudinal clinical outcomes.

| Outcomes | CN | LBD-nlCog | LBD-abnlCog | AD |
| --- | --- | --- | --- | --- |
| | $\beta$ estimate (SE) | | | |
| **MoCA** | -0.17 (0.48) | 1.35 (1.52) | -2.09 (1.83) | **-5.55 (1.26)\*\*\*** |
| **CDR-SB** | 0.10 (0.22) | -0.15 (0.67) | **1.97 (0.80)\*** | **2.65 (0.40)\*\*\*** |
| **Memory** | -0.08 (0.12) | 0.18 (0.34) | **-1.31 (0.43)\*\*** | -0.50 (0.42) |
| **Executive function** | -0.08 (0.10) | 0.09 (0.32) | -0.70 (0.38) | -0.57 (0.37) |
| **Attention/WM/PS** | **-0.16 (0.07)\*** | -0.11 (0.22) | -0.54 (0.30) | 0.07 (0.30) |
| **Visuospatial function** | -0.24 (0.16) | 0.15 (0.46) | 0.50 (0.68) | 0.02 (0.60) |
| **Language** | -0.14 (0.21) | -0.00 (0.71) | -0.39 (0.83) | **-1.54 (0.67)\*** |

\* p<0.05 \*\*p<0.01 \*\*\* p<0.001
Abbreviations: AD: Alzheimer's disease, CDR-SB: Clinical Dementia Rating Sum of Boxes, CN: cognitively normal, LBD-nlCog: LBD with normal cognition, LBD-abnlCog: LBD with abnormal cognition, MoCA: Montreal Cognitive Assessment, SE: standard error, PS: processing speed, WM: working memory.

### 1. 6. Plasma pTau181 as a predictive biomarker for conversion to MCI or dementia in LBD patients with normal cognition

We categorized participants from the CN and LBD-nlCog groups into those with normal and abnormal plasma pTau181 levels, using the cut-point of 1.71 pg/mL determined by the Youden J Index method to distinguish LBD-nlCog Aβ+ from Aβ-individuals. Then, we identified participants who progressed to cognitive impairment using longitudinal data from consensus meetings (longitudinal data available: 173 out of 274 CN, and 61 out of 94 LBD-nlCog participants). From a total of 234 participants with longitudinal data over a 6-years follow-up period, 25 (11%) converted to cognitive impairment over a period of 0.73 to 4.87 years; 14 (8%) CN participants (time to conversion: 0.8-4.9 years, median: 1.81 years), and 11 (18%) LBD-nlCog participants (time to conversion: 0.7-3.2 years, median: 1.99 years).

We found that individuals in the LBD-nlCog group with abnormal baseline plasma pTau181 levels had a significantly higher risk of developing cognitive impairment than those in the CN group with normal levels (reference group) (HR: 5.2, 95% CI: 1.6-17.2, p<0.01). Interestingly, there was a trend towards significance in the risk of developing cognitive impairment between LBD-nlCog group with abnormal plasma pTau181 levels and the LBD-nlCog group with normal levels (HR: 2.98, 95% CI: 1.0-9.3, p=0.061). By contrast, there was no significant difference in the risk of developing cognitive impairment between the CN group with abnormal plasma pTau181 levels and LB-nlCog group with normal levels when compared to the CN group with normal levels **(Figure 2)**.

**Figure 2:**
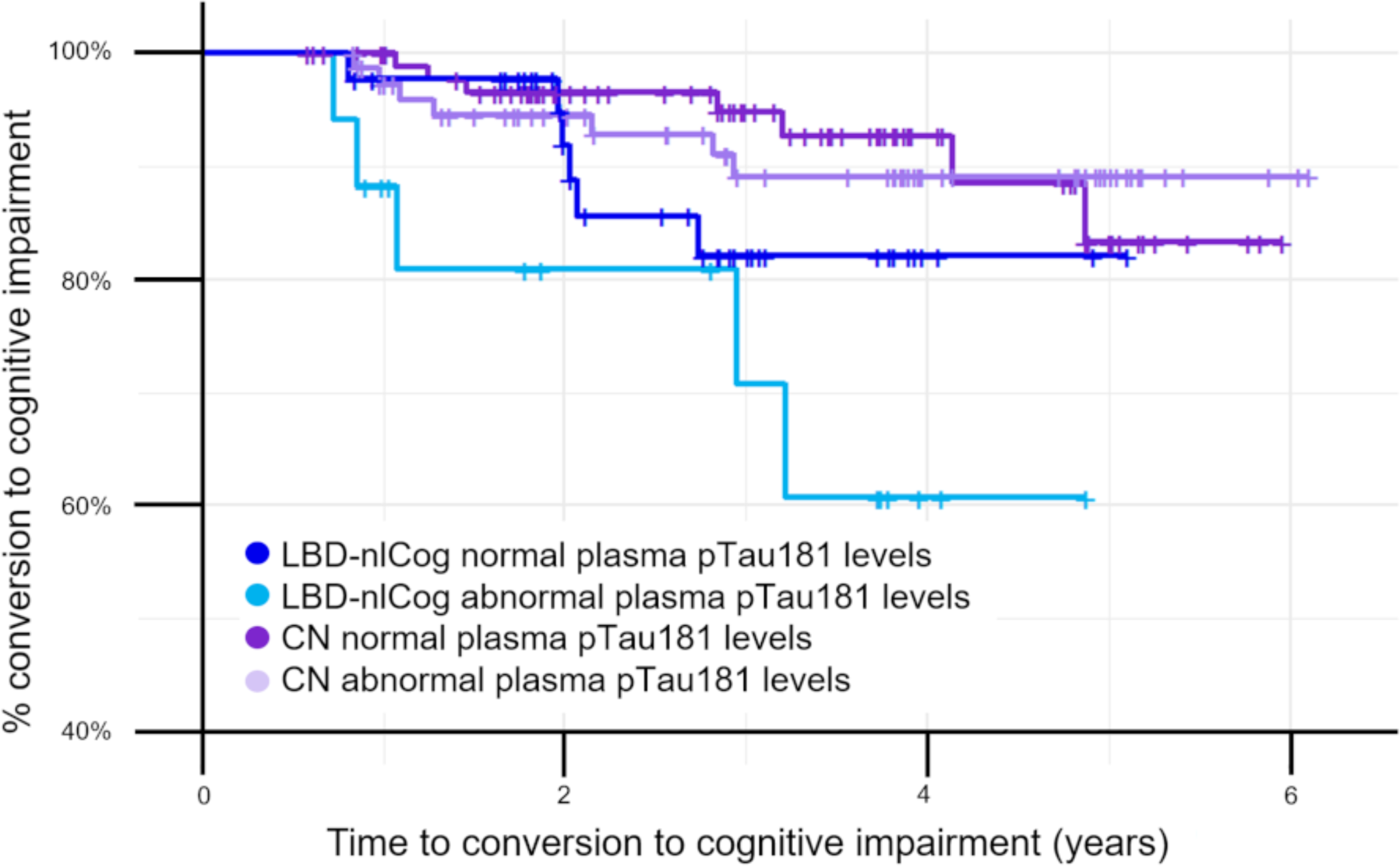
Survival curves of conversion to cognitive impairment (MCI or dementia) in CN and LBD-nlCog participants with normal and abnormal plasma pTau181 levels. LBD-nlCog with normal plasma pTau181 levels (N= 44, dark blue line), LBD-nlCog with abnormal plasma pTau181 levels (N= 17, light blue line), CN with normal plasma pTau181 levels (N= 95, purple line), CN with abnormal plasma pTau181 levels (N= 78, light purple line). Normal plasma pTau181 levels were defined as < 1.71 pg/mL, while abnormal levels were ≥ 1.71 pg/mL, using a cut-point determined by the Youden J Index method to distinguish LBD-nlCog Aβ+ from Aβ-individuals. Abbreviations: Aβ: amyloid-β, CN: cognitively normal, LBD-nlCog: Lewy body disease with normal cognition, pTau181: phosphorylated-Tau181.

## DISCUSSION

In this study, we evaluated plasma pTau181 as a potential diagnostic biomarker for ADNC and amyloidosis, as well as a prognostic, monitoring, and susceptibility/risk biomarker in LBD participants. Our findings revealed that plasma pTau181 in LBD not only detects concurrent ADNC and amyloidosis but is also associated with baseline and longitudinal clinical outcomes in LBD participants with cognitive impairment, as well as with progression to cognitive impairment during follow-up in LBD participants with normal cognition at baseline. These results are relevant for clinical practice, as they indicate the potential use of plasma pTau181 for identifying LBD patients with worse prognosis. This information can help guide clinical care and provide valuable insights for patients and their caregivers. Moreover, the potential use of plasma pTau181 in LBD drug development is promising, as it may facilitate the enrichment of clinical trials with participants more likely to progress, allow for stratification based on the presence or absence of concomitant ADNC or amyloidosis, and may enable the detection of treatment response^40,41^.

In line with previous studies, we demonstrated that in LBD participants with cognitive impairment, plasma pTau181 can detect concurrent ADNC and amyloidosis with relatively high accuracy^14–16^, and that elevated plasma pTau181 levels are associated with worse baseline global cognition^14,42^. Moreover, to our knowledge, this is the first report of plasma pTau181 detecting concurrent amyloidosis in LBD patients with normal cognition (i.e. PD with normal cognition), and its association with a measure of daily functioning (CDR-SB) in LBD participants with MCI or dementia. Notably, our findings also revealed high negative predictive values for plasma pTau181 in detecting concurrent ADNC in LBD participants with cognitive impairment, and amyloidosis in LBD participants with normal cognition. This suggests that normal levels of plasma pTau181 could be used in clinic practice to rule out the presence of concurrent ADNC and amyloidosis in these groups of patients. In contrast, though the positive predictive values do not unambiguously confirm the presence of concurrent ADNC and amyloidosis in LBD, they may imply the need for more sensitive tests like CSF AD biomarkers or PET neuroimaging, or a repeated plasma pTau181 test during a follow-up visit. Previous studies using CSF AD biomarkers and PET neuroimaging have allowed detection of concurrent ADNC in LBD participants, mirroring the findings from autopsy studies^43–45^. Collectively, our findings underscore the potential of plasma pTau181 as non-invasive and cost-effective diagnostic tool for detecting concurrent ADNC and amyloidosis in LBD patients, serving as a preliminary screening test before resorting to more invasive and expensive biomarker assessments. Additionally, our findings raise questions about using plasma pTau181 to distinguish AD from LBD patients, as those with LBD may also exhibit concurrent ADNC. This emphasizes the importance of considering co-pathologies when interpreting biomarker results.

Our findings indicate that plasma pTau181 is a promising prognostic biomarker in LBD patients with cognitive impairment, as we demonstrated that higher baseline levels might be associated with a more rapid decline in daily functioning over time. Identifying people with LBD and cognitive impairment, who are at higher risk for faster functional decline can inform patient management, and facilitate the stratification and enrichment of clinical trials. For example, a recent clinical trial investigating neflamapimod in DLB patients revealed that participants with low baseline plasma pTau181 levels in the treatment group exhibited improvement in all endpoints when compared to the placebo group^46^.

In contrast to previous research, we did not find an association with baseline plasma pTau181 levels and subsequent decline in global cognition. Previous studies have reported the prognostic value of plasma pTau181 in detecting LBD with a faster global cognitive decline at the dementia stage^14^ but not at the MCI stage^16^. A possible explanation is that we combined both MCI and dementia stages and used a different outcome measure for global cognition (MoCA).

Importantly, our study introduces the potential use of plasma pTau181 as a monitoring biomarker in LBD for the first time. Individuals with MCI and dementia in the context of LBD, alongside ADNC or amyloidosis alone, tend to experience faster disease progression with greater cognitive impairment that those without these co-pathologies^6,42^. The ability to repeatedly measure biomarkers using blood samples now provides a means to identify patients likely to have a more aggressive disease course. Such information could potentially be used in clinical trials to detect treatment response. We observed that changes in plasma pTau181 levels over 3 years predict a more rapid decline in daily functioning and memory performance. In contrast to our results, a recent study did not find an association between the change in plasma pTau181 levels and cognitive decline in MCI-LB^16^. This discrepancy in findings may be attributed to the differences in the assessment tools, and the composition of the diagnostic groups. Future studies should extend our results to prodromal LBD participants to determine if changes in plasma pTau181 predict more rapid cognitive decline at this earlier disease stage.

Another relevant discovery in our study was the ability of plasma pTau181 to identify LBD participants with normal cognition (i.e., PD with normal cognition) at an increased risk of progressing to MCI or dementia, which is aligned with previous work analyzing longitudinal AD CSF biomarkers studies in PD^47^. When comparing LBD patients with normal cognition and abnormal baseline plasma pTau181 levels to the CN group with normal levels, we found an evident increase in risk. Of particular importance was the observed trend indicating a nearly threefold higher risk of progression to cognitive impairment in LBD participants with normal cognition and abnormal levels of plasma pTau181, compared to those with normal levels. It’s important to note that our study may have been underpowered to detect a stronger association, given the limited sample size of 17 subjects in the LBD-nlCog group with normal cognition and abnormal baseline levels of plasma pTau181. These results might suggest that plasma pTau181 can serve as a blood-based susceptibility/risk biomarker in PD, aiding in the identification of patients at higher risk of developing cognitive impairment in clinical and research settings.

Another study by Pagobarraga et al obtained different findings; they did not observe a predictive relationship between plasma pTau181 levels and the progression to dementia^17^. Differences in the analytical approach might explain the divergent results. They investigated the transition to dementia in a combined group of PD participants with normal cognition and MCI, while we analyzed the transition to cognitive impairment in participants with normal cognition at baseline. We hypothesized that the stratification of LBD participants based on cognitive status (i.e., normal cognition vs cognitive impairment) is critical when exploring the link between concurrent amyloidosis and cognitive decline in LBD. This distinction is grounded in the potential synergistic effect of α-synuclein, amyloid and tau deposition in the neocortex, which may contribute to cognitive deficits in LBD^48,49^.

Consistent with previous research, we observed that plasma pTau181 levels at baseline were not associated with cognitive performance or daily functioning at either the initial assessment or over time in LBD participants with normal cognition^50,51^. Importantly, changes in plasma pTau181 levels over time did not predict changes in domain-specific cognitive test scores or daily functioning either.

Our study has strengths and limitations. Among its strengths are a large sample size, the availability of longitudinal data, and the inclusion of patients across the entire LBD continuum, ranging from participants with normal cognition to those with dementia. However, there are some limitations. Firstly, this study was conducted in a single center (Stanford University), emphasizing the need for validation in an independent cohort. Additionally, while our study focused on plasma pTau181, other plasma pTau isoforms such as pTau217 and pTau231 may provide enhanced sensitivity in detecting ADNC in LBD^14,15^. Another limitation is the lack of diversity in our sample population, which primarily consisted of a convenience sample of non-Hispanic whites, potentially limiting the generalizability of our findings. Lastly, although we carefully excluded patients with known renal or hepatic disorders, future studies should investigate the potential impact of comorbidities on plasma biomarkers in LBD.

In summary, our study emphasizes the importance of studying the presence of co-pathologies in neurodegenerative diseases. We elucidated the practical implications of finding ADNC and amyloidosis in LBD patients, revealing the association of co-pathology with negative clinical outcomes. This observation suggests that treatment effects may vary among patients with and without mixed pathologies, highlighting the need to identify such individuals for participant selection and stratification in clinical trials. Beyond research, these findings have relevant applications for clinical practice. They can guide patient management, inform patients and their caregivers about prognosis, and help allocate healthcare resources more effectively. Thus, recognizing and understanding the presence of both ADNC and amyloidosis in LBD represents a step toward recognizing a biological etiology underlying heterogeneity in clinical progression, optimizing patient care, and overall advancing our understanding of these complex neurodegenerative conditions.

## Data Availability

Anonymized data supporting the conclusions of the current study is available to qualified researchers upon reasonable request, but this will be contingent upon the authors ability to secure Stanford University's institutional review board approval, and upon both parties signing a data use agreement.

