## Supplementary material for "Plasma pTau181 reveals a pathological signature that predicts cognitive outcomes in Lewy body disease"

**Supplementary Figure 1: Selection of research participants.**

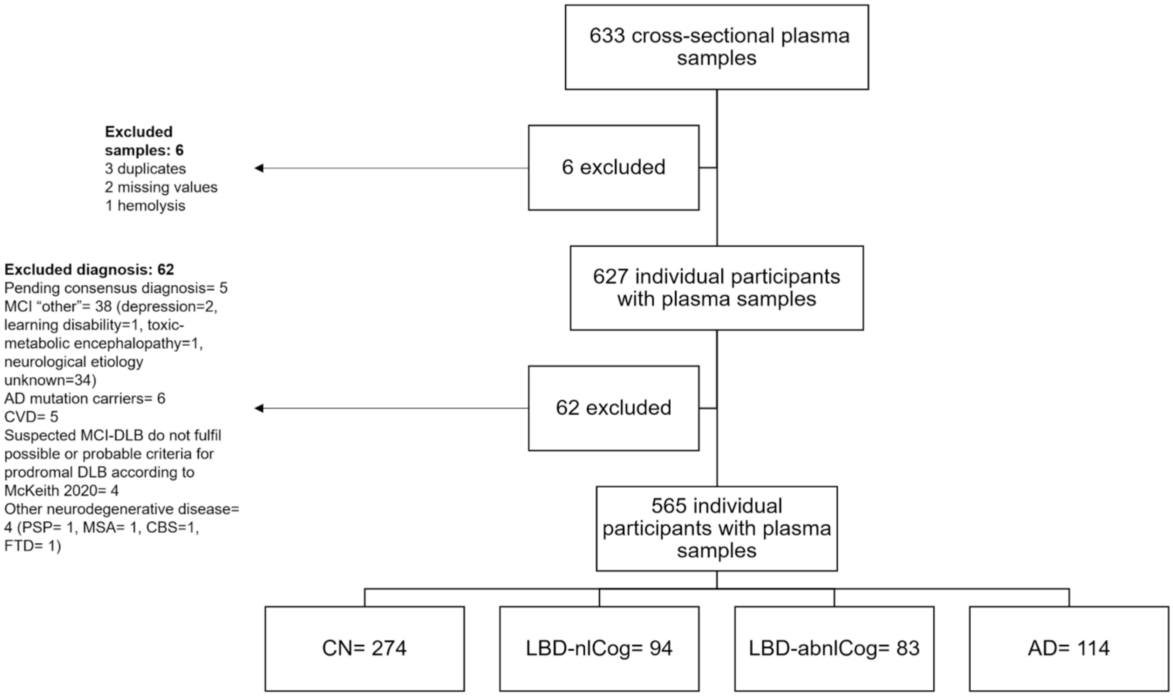


Abbreviations: AD: Alzheimer's disease, CBS: corticobasal syndrome, CSF: cerebrospinal fluid, CN: cognitively normal, CVD: cerebrovascular disease, DLB: dementia with Lewy bodies, FTD: frontotemporal dementia, LBD-nlCog: LBD with normal cognition, LBD-abnlCog: LBD with abnormal cognition, MCI: mild cognitive impairment, MSA: multiple system atrophy, PET: positron emission tomography, PSP: progressive supranuclear palsy.

**Supplementary Figure 2: Associations between 3-years change in plasma pTau181 levels and longitudinal clinical outcomes.**

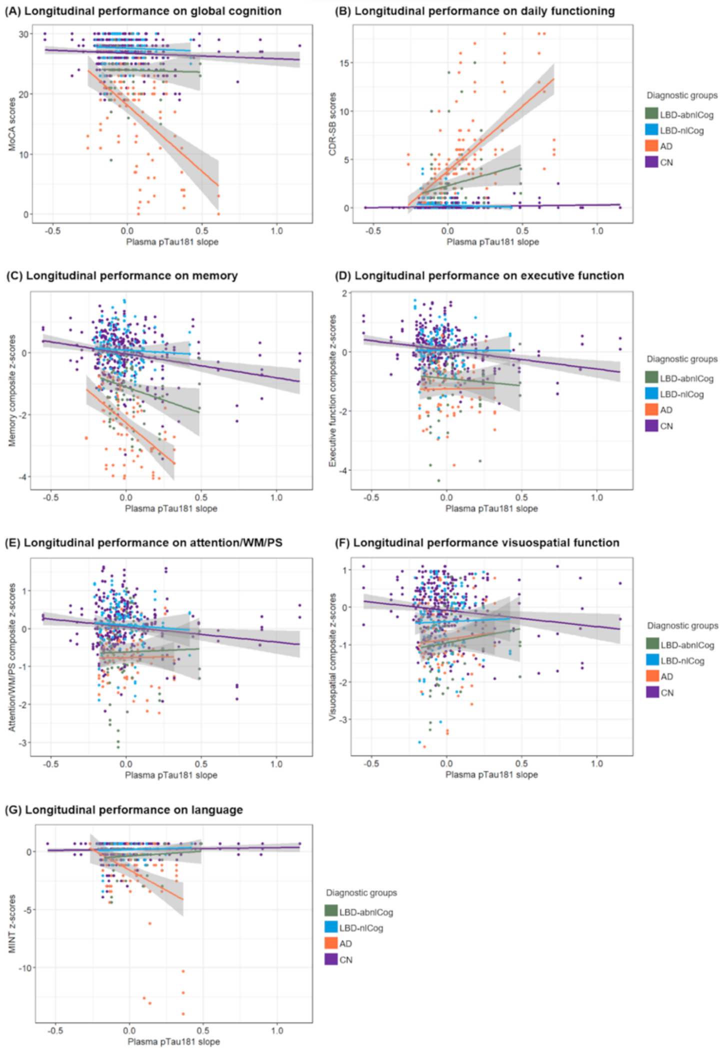


Each line represents the association between the longitudinal performance on clinical outcomes with the change in plasma pTau181 levels over 3 years for the respective diagnostic group: LBD-abnlCog (green line), LBD-nlCog (blue line), AD (orange line), and CN (purple line). Trajectories were calculated using linear mixed effects models with the slope of plasma pTau181 levels over 3 years by time as predictor, as well as interaction terms between covariates and time: age and sex, and year of education for cognitive outcomes. All models incorporated random intercepts and slopes. **A)** Longitudinal performance on global cognition (MoCA) **B)** Longitudinal performance on daily functioning (CDR-SB) **C-G)** Longitudinal performance on neuropsychological assessment. Z-scores for each neuropsychological test were calculated based on the means and standard deviations from the baseline visit of all CN participants. These z-scores were then used to create composite scores for specific domains by averaging the relevant z-scores. Plasma pTau181 levels were log-transformed. **Supplementary Table 2** shows the number of participants with 3-years change in plasma pTau181 levels and clinical longitudinal data.

Abbreviations: AD: Alzheimer's disease, CDR-SB: Clinical Dementia Rating Sum of Boxes, CN: cognitively normal, LBD-abnlCog: Lewy body disease with abnormal cognition, LBD-nlCog: Lewy body disease with normal cognition, MINT: multilingual naming test, MoCA: Montreal Cognitive Assessment, pTau181: phosphorylated-Tau181.

**Supplementary Table 1: Number of participants with baseline plasma pTau181 levels and clinical longitudinal data**

| Variables | Overall | Baseline | Year 1 | Year 2 | Year 3 |
|---|---|---|---|---|---|
| **MoCA** | 307 | 303 | 286 | 69 | 110 |
| **CDR-SB** | 351 | 343 | 349 | 299 | 225 |
| **Memory** | 264 | 255 | 241 | 62 | 115 |
| **Executive function** | 249 | 241 | 224 | 57 | 92 |
| **Attention/WM/PS** | 249 | 241 | 224 | 55 | 89 |
| **Visuospatial function** | 257 | 238 | 233 | 58 | 111 |
| **Language** | 257 | 255 | 232 | 65 | 94 |

Abbreviations: CDR-SB: Clinical Dementia Rating Sum of Boxes, MoCA: Montreal Cognitive Assessment, PS: processing speed, pTau181: phosphorylated-Tau181, WM: working memory.

**Supplementary Table 2: Number of participants with 3-years change in pTau181 levels and clinical longitudinal data**

| Variables | Overall | Baseline | Year 1 | Year 2 | Year 3 |
|---|---|---|---|---|---|
| **MoCA** | 231 | 227 | 223 | 54 | 100 |
| **CDR-SB** | 244 | 238 | 243 | 229 | 205 |
| **Memory** | 219 | 208 | 214 | 49 | 103 |
| **Executive function** | 216 | 210 | 209 | 43 | 82 |
| **Attention/WM/PS** | 219 | 211 | 212 | 41 | 81 |
| **Visuospatial function** | 211 | 210 | 205 | 58 | 135 |
| **Language** | 223 | 220 | 217 | 51 | 84 |

Abbreviations: CDR-SB: Clinical Dementia Rating Sum of Boxes, MoCA: Montreal Cognitive Assessment, PS: processing speed, pTau181: phosphorylated-Tau181, WM: working memory.

**Supplementary Table 3: Characteristics of research participants with CSF biomarkers or amyloid PET**

| Variable | CN (n=115) | LBD-nlCog (n=47) | LBD-abnlCog (n=47) | AD (n=36) | Total (n=245) | *p* values |
|---|---|---|---|---|---|---|
| **Age, years** | | | | | | |
| Mean (SD) | 68.66 (7.91) | 67.77 (7.28) | 69.77 (6.82) | 71.00 (8.51) | 69.04 (7.71) | 0.231 |
| Range | 51-92 | 50-80 | 57-85 | 55-91 | 50-92 | |
| **Sex** | | | | | | |
| n (%) males | 52 (45.2) | 27 (57.4) | 33 (70.2) | 17 (47.2) | 129 (52.7) | 0.026 |
| **Education, years** | | | | | | |
| Median (range) | 17 (5-20) | 16 (12-20) | 18 (12-20) | 16 (8-20) | 16 (5-20) | 0.149 |
| **Race,** n (%) | | | | | | |
| White | 103 (89.6) | 41 (87.2) | 45 (95.7) | 31 (86.1) | 220 (89.8) | 0.237 |
| Black or African American | 1 (0.9) | 0 | 0 | 1 (2.8) | 2 (0.8) | |
| American Indian or Alaska Native | 0 | 1 (2.1) | 0 | 2 (5.6) | 3 (1.2) | |
| Asian | 9 (7.8) | 5 (10.6) | 2 (4.3) | 1 (2.8) | 17 (6.9) | |
| Other | 2 (1.7) | 0 | 0 | 1 (2.8) | 3 (1.2) | |
| **Ethnicity**, n (%) | | | | | | |
| Not Hispanic or Latino | 99 (86.1) | 47 (100) | 44 (93.6) | 33 (91.7) | 223 (91.0) | 0.037 |
| Hispanic or Latino | 16 (13.9) | 0 | 3 (6.4) | 3 (8.3) | 22 (9.0) | |
| **Years since symptom onset** | | | | | | |
| Median (range) | - | 8 (1-29)[d] | 7 (1-24) | 4 (0-20)[b] | 6 (0-29) | 0.028 |
| **MoCA** | | | | | | |
| Median (range) | 27 (12-30)[c,d] | 28 (20-30)[c,d] | 23 (3-28)[a,b] | 16 (1-27)[a,b] | 26 (1-30) | <0.001 |
| **CDR-SB** | | | | | | |
| Median (range) | 0 (0-3.5)[c,d] | 0 (0-1)[c,d] | 1.5 (0-11)[a,b] | 4 (0-17)[a,b] | 0 (0-17) | <0.001 |
| **MDS-UPDRS Part III Off** | | | | | | |
| Median (SD) | - | 33.74 (16.01)[d] | 32.92 (16.63)[d] | 8.73 (8.70)[b,c] | 16.96 (18.06) | <0.001 |
| **LEDD, mg/d** | | | | | | |
| Median (range) | - | 625 (100-1580) | 600 (260-1547) | - | 612.50 (100-1580) | 0.932 |
| **APOE ε4 carrier,** n (%) | 35 (30.4)[d] | 14 (29.8)[d] | 13 (27.7)[d] | 21 (58.3)[a,b,c] | 83 (33.9) | <0.006 |
| Missing, n (%) | 15 (13.0) | 3 (6.4) | 5 (10.6) | 4 (11.1) | 27 (11.0) | |
| **Plasma pTau181 levels, pg/mL** | | | | | | |
| Mean (SD) | 1.60 (0.62)[c,d] | 1.56 (0.59)[c,d] | 2.09 (1.38)[a,b,d] | 3.00 (1.31)[a,b,c] | 1.89 (1.05) | <0.001 |
| Median | 1.44 | 1.44 | 1.78 | 2.5 | 1.57 | |
| Range | 0.73-4.63 | 0.93-4.39 | 1.03-10.43 | 1.32-6.13 | 0.73-10.43 | |
| **AD CSF biomarkers** | | | | | | |
| **Aβ42, pg/mL** | | | | | | |
| Mean (SD) | 1023.26 (415.50)[c,d] | 922.98 (362.43)[c,d] | 695.94 (363.85)[a,b] | 686.50 (274.29)[a,b] | 894.38 (404.67) | <0.001 |
| **Total tau, pg/mL** | | | | | | |
| Mean (SD) | 321.63 (239.88)[d] | 260.58 (140.36)[c,d] | 414.61 (284.58)[b,d] | 683.61 (401.42)[a,b,c] | 378.86 (292.81) | <0.001 |

| | | | | | | |
|---|---|---|---|---|---|---|
| **pTau181, pg/mL**<br>Mean (SD) | 43.87 (32.54)[d] | 32.59 (11.49)[c,d] | 50.85 (31.29)[b,d] | 105.47 (75.83)[a,b,c] | 51.83 (44.82) | <0.001 |
| **Aβ42/40 ratio**<br>Mean (SD) | 0.11 (0.03)[d] | 0.12 (0.02)[c,d] | 0.10 (0.04)[b] | 0.08 (0.03)[a,b] | 0.10 (0.31) | <0.001 |
| **pTau181/Aβ42 ratio**<br>Mean (SD) | 0.05 (0.06)[c,d] | 0.039 (0.18)[d] | 0.09 (0.06)[a] | 0.18 (0.13)[a,b] | 0.75 (0.82) | <0.001 |
| **Amyloid PET positive.** N (%) | 8 (23.5)[d] | 1 (7.7)[d] | 4 (36.4)[d] | 12 (80.0)[a,b,c] | 25 (34.2) | <0.001 |
| **ADNC present.**<br>N (%)<br>Missing, n (%) | 5 (4.3)[c,d]<br>0 | 0[d]<br>1 (2.1) | 9 (19.1)[a]<br>1 (2.1) | 19 (52.8)[a,b]<br>0 | 33 (13.5)<br>2 (0.8) | <0.001 |
| **Amyloidosis.**<br>N (%) | 32 (27.8)[c,d] | 6 (12.8)[d] | 24 (51.1)[a] | 29 (80.6)[a,b] | 91 (37.1) | <0.001 |

*p* values (uncorrected) were calculated using ANOVA, chi-square, and Kruskal-Wallis tests as appropriate. Pair-wise comparisons were corrected for multiple comparisons using the Bonferroni correction. [a]p <0.05 compared to CN, [b]p <0.05 compared to LBD with normal cognition, [c]p <0.05 compared to LBD with abnormal cognition, [d]p <0.05 compared to AD.

Abbreviations: Aβ: amyloid-β, AD: Alzheimer's disease, ADNC: Alzheimer's Disease Neuropathologic Change, APOE: apolipoprotein E, CDR-SB: Clinical Dementia Rating Sum of Boxes, CN: cognitively normal, CSF: cerebrospinal fluid, LBD-nlCog: LBD with normal cognition, LBD-abnlCog: LBD with abnormal cognition, LEDD: levodopa equivalent daily dose, MDS-UPDRS: United Parkinson's Disease Rating Scale, Movement Disorder Society revision; MoCA: Montreal Cognitive Assessment, PET: positron emision tomography, pTau181: phosphorylated-Tau181, SD: standard deviation. * Years since symptom onset was calculated from age of symptom onset to age at plasma collection.
